# Differential DNA methylation landscape in skin fibroblasts from African Americans with systemic sclerosis

**DOI:** 10.1101/2020.08.12.20173773

**Authors:** DeAnna Baker Frost, Willian da Silveira, E. Starr Hazard, Ilia Atanelishvili, Robert C. Wilson, Jonathan Flume, Kayleigh L. Day, James C. Oates, Galina S. Bogatkevich, Carol Feghali-Bostwick, Gary Hardiman, Paula S. Ramos

**Author notes:** **Correspondence to** Dr. Paula S. Ramos, Division of Rheumatology & Immunology, Department of Medicine, Medical University of South Carolina, 96 Jonathan Lucas Street, MSC637, Charleston, SC 29425.

## Abstract

**Objective:** The etiology and reasons underlying the ethnic disparities in systemic sclerosis (SSc) remain unknown. African Americans are disproportionally affected by SSc, yet underrepresented in research. The aim of this study was to comprehensively investigate the association of DNA methylation levels with SSc in dermal fibroblasts from patients of African ancestry.

**Methods:** Reduced representation bisulfite sequencing (RRBS) was performed on primary cultured dermal fibroblasts from 15 SSc patients and 15 controls of African ancestry, and over 3.8 million CpG sites were tested for differential methylation patterns between cases and controls. Gene set enrichment (GSEA) and gene ontology (GO) analyses were computed to elucidate the underlying biological processes. Quantitative PCR (qPCR) was performed to assess correlations between DNA methylation changes and gene expression levels of top candidate genes.

**Results:** Skin fibroblasts from African American patients exhibited widespread reduced DNA methylation. Differentially methylated CpG sites were most enriched in introns and intergenic regions, while depleted in 5’ UTR, promoters, and CpG islands. Seventeen genes and eleven promoters showed significant differential methylation, mostly in non-coding RNA genes and pseudogenes. GSEA and GO enrichment analysis revealed enrichment of immune, metabolism, cell development, and cell signaling pathways, including those related to interferon signaling and mesenchymal differentiation. The hypomethylation of *DLX5 and TMEM140* was accompanied by these genes’ overexpression, while for the lncRNA *MGC12916*, it was accompanied by its under-expression in patients.

**Conclusion:** These data show that differential methylation occurs in dermal fibroblasts from African American patients with SSc and identifies novel coding and non-coding genes.

## INTRODUCTION

Systemic sclerosis (SSc or scleroderma) is a multisystem, connective tissue disease characterized by cutaneous and visceral fibrosis, immune dysregulation, and vasculopathy. Patients are commonly classified into two main subsets: limited cutaneous SSc (lcSSc) and diffuse cutaneous SSc (dcSSc), with dcSSc having a worse prognosis.^1^ Relative to individuals of European ancestry, individuals of African ancestry are more likely to develop SSc, to be diagnosed with dcSSc, and to experience higher disease severity, greater morbidity, reduced survival, and earlier death.^2-9^ This higher disease burden in African Americans is not fully explained by differences in socioeconomic status or access to health care.^9,10^

The etiology of SSc and the factors underlying its ethnic disparities remain elusive. Genetic and epigenetic studies conducted mostly in individuals of European ancestry uncovered multiple loci associated with SSc.^11^ A role for DNA methylation in SSc is supported by a X chromosome gene methylation analysis of peripheral blood mononuclear cells,^12^ quantification of global methylation in whole blood,^13^ as well as genome-wide DNA methylation analyses of dermal fibroblasts,^14^ whole blood,^15^ and CD4+ T cells.^16^ Different ancestral populations exhibit DNA methylation differences^17-24^ that are partially explained by their distinct genetic ancestry, thus environmental factors not captured by genetic ancestry are significant contributors to variation in methylation.^19^

In order to understand the pathogenesis of SSc in patients of African ancestry, we assessed DNA methylation profiles of dermal fibroblasts from African American patients and controls by RRBS, which has high sensitivity and specificity to detect changes in DNA methylation in genes, promoters, CpG islands, and repetitive regions.^25,26^ We then integrated the data with gene expression of the top differentially methylated genes from the same subjects. This study is the first to unveil the genome-wide patterns of differential methylation in skin fibroblasts from African American patients with SSc.

## METHODS

### Subjects

A total of 15 SSc cases and 15 healthy controls were recruited for this study. All participants were self-reported African American and patients met the 2013 ACR/EULAR classification criteria for SSc. Cases and controls were age-balanced within 5 years. This study was approved by the IRB at the Medical University of South Carolina.

### Primary dermal fibroblast isolation and culture

Primary dermal fibroblasts were isolated and cultured as described.^27^ Cells were cultured for 3 passages, then DNA and RNA were isolated using DNeasy and RNeasy kits (Qiagen, Germantown, MD) following the manufacturer’s protocols.

### Reduced representation bisulfite sequencing (RRBS)

RRBS was performed using the Ovation® RRBS Methyl-Seq System 1–16 (NuGEN Technologies, Inc., San Carlos, CA) following the manufacture’s procedure. We generated DNA methylation data for over 5 million CpGs in each sample and between 10x to 40x coverage in CpG sites.

### Genome-wide DNA methylation data analysis

Alignment and methylation calling were performed using Bismarck v0.16.3 and the GRCh37/hg19 reference genome.^28^ Data was filtered, normalized, and analyzed with RnBeads v1.6.1.^29^ Differential methylation analysis was conducted at CpG, promoter, and gene levels (including RNA, pseudo-,and protein-coding genes).^29^ Genes and promoters were defined by Ensembl and CpG islands were defined as on the CpG island track of the UCSC Genome Browser. As implemented in RnBeads, CpG site *p*-values were computed using the linear models in the limma package. Genes, promoters and CpG islands were ranked based on the RnBeads’ Combined Score approach.^29^

### Genomic annotation enrichment analysis

To annotate the position of each CpG to the corresponding genomic location, the *annotatePeaks.pl* program of Homer^30^ was used. CpGs were annotated to promoter, transcription termination site (TTS), exon, intron, 5’ UTR exon, 3’ UTR exon, intergenic, CpG island, repeat elements, and other detailed annotations. To investigate the distribution of differentially methylated CpGs (DMC) in different genomic locations, all CpGs that met an FDR-adjusted *ρ*-value < 0.4 were used to compare their localization in different genomic locations as provided by of Homer’s annotations^30^. Odds ratio (OR), 95% confidence intervals (CI), and *ρ*-values were computed against the general distribution of the 3,870,251 CpGs of our dataset using GraphPad Prism.

### Gene set enrichment analysis (GSEA)

GSEA was performed to determine whether a priori defined sets of genes (e.g. pathways) are significantly enriched in the list of genes ranked by their correlation with the disease. The full ranked lists of genes and promoters generated by RnBeads’ Combined Score approach^29^ were used as input to GSEA Desktop v3.0.^31,32^ The genes were ranked by their differential methylation between cases and controls (hyper- and hypomethylated), and the Reactome Pathway Knowledgebase (https://reactome.org)^33^ was used as the gene set. An enrichment score statistic represents the enrichment of Reactome pathways in the genes that are hyper- or hypomethylated in patients, and the significance of the pathway enrichment score is estimated by an empirical phenotype-based permutation test procedure.^31,32^ The threshold for statistical significance was defined as FDR ≤ 0.25.^31,32^

### Gene ontology (GO) enrichment analysis

Enrichment analysis for GO terms associated with the top-ranking differentially methylated genes and promoters was performed using RnBeads v1.6.1.^29^ GO enrichment analysis of biological process (BP) was conducted on each of the 100 hypo- and hypermethylated genes and promoters. Enrichment of GO BP terms associated with the top ranking genes and promoters was determined by a hypergeometric test implemented in RnBeads.^29^

### Gene expression analysis

cDNA was prepared using Superscript IV First Strand synthesis system (ThermoFisher, Waltham, MA) from 1 μg of isolated RNA. qPCR was performed using Taqman Real-Time PCR master mix (Applied Biosystems, Foster City, CA). All samples were run in duplicate using Applied Biosystems Real-Time PCR System and analyzed using StepOne Plus Applied Biosystems software. Gene quantification cycle values were normalized to B2M expression using the ΔΔCT method to obtain relative cell equivalents. All primers were purchased from ThermoFisher Scientific (Waltham, MA). Statistical significance was determined using the Mann-Whitney test and defined as *p*-value ≤ 0.05.

## RESULTS

### Subject demographics

The clinical and demographic characteristics of the volunteer African ancestry SSc patients and healthy controls are summarized in table 1. Most patients were female with dcSSc and no concomitant rheumatic disease.

**Table 1.**
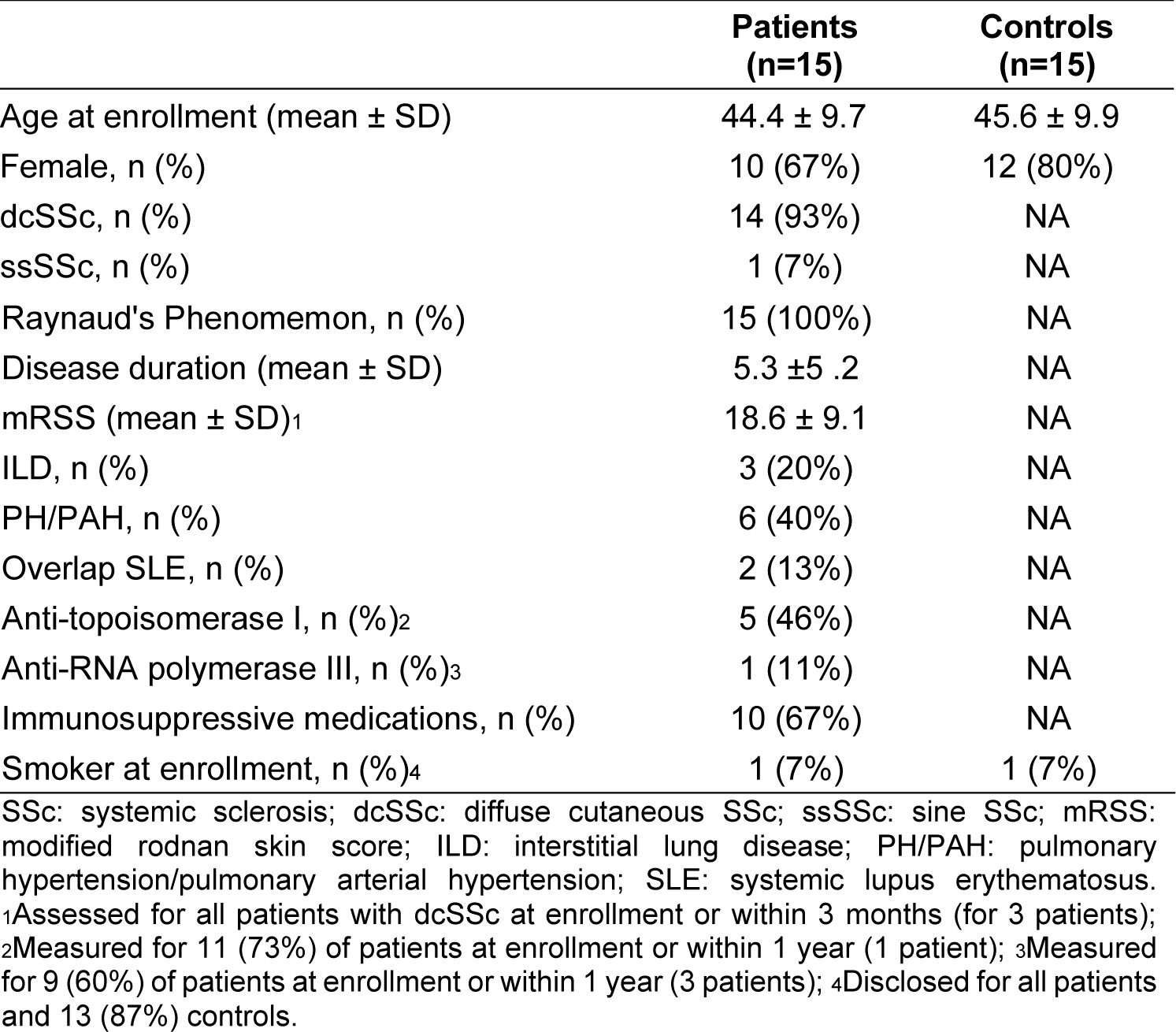
Demographic and Clinical Characteristics of Study Participants

|  | Patients<br>(n=15) | Controls<br>(n=15) |
| --- | --- | --- |
| Age at enrollment (mean ± SD) | 44.4 ± 9.7 | 45.6 ± 9.9 |
| Female, n (%) | 10 (67%) | 12 (80%) |
| dcSSc, n (%) | 14 (93%) | NA |
| ssSSc, n (%) | 1 (7%) | NA |
| Raynaud's Phenomenon, n (%) | 15 (100%) | NA |
| Disease duration (mean ± SD) | 5.3 ± 5.2 | NA |
| mRSS (mean ± SD)<sup>1</sup> | 18.6 ± 9.1 | NA |
| ILD, n (%) | 3 (20%) | NA |
| PH/PAH, n (%) | 6 (40%) | NA |
| Overlap SLE, n (%) | 2 (13%) | NA |
| Anti-topoisomerase I, n (%)<sup>2</sup> | 5 (46%) | NA |
| Anti-RNA polymerase III, n (%)<sup>3</sup> | 1 (11%) | NA |
| Immunosuppressive medications, n (%) | 10 (67%) | NA |
| Smoker at enrollment, n (%)<sup>4</sup> | 1 (7%) | 1 (7%) |

SSc: systemic sclerosis; dcSSc: diffuse cutaneous SSc; ssSSc: sine SSc; mRSS: modified rodnan skin score; ILD: interstitial lung disease; PH/PAH: pulmonary hypertension/pulmonary arterial hypertension; SLE: systemic lupus erythematosus. <sup>1</sup>Assessed for all patients with dcSSc at enrollment or within 3 months (for 3 patients); <sup>2</sup>Measured for 11 (73%) of patients at enrollment or within 1 year (1 patient); <sup>3</sup>Measured for 9 (60%) of patients at enrollment or within 1 year (3 patients); <sup>4</sup>Disclosed for all patients and 13 (87%) controls.

### Differentially methylated sites and genes

Over 3.8 million CpG sites were tested for differential methylation between SSc cases and controls (supplementary figure S1). Patients exhibited widespread hypomethylation throughout the genome, with over 85% of CpGs that met an FDR-adjusted *p*-value < 0.4 showing decreased methylation in the cultured skin fibroblasts from the patients compared to the controls. The rationale for the FDR setting was guided by the desire to perform a systems level analysis and include as many CpGs sites as possible. Among the 1,180 DMCs that met an adjusted *p*-value < 0.4, there was an overrepresentation of DMCs in introns (OR = 1.7, *p* < 0.0001), intergenic regions (OR = 1.5, *p* < 0.0001), TTS (OR = 1.5, *p* = 0.007), and short interspersed nuclear elements (SINE) (OR = 1.2, *p* = 0.003) (figure 1 and supplementary table S1). Notably, there was a depletion of DMCs in 5’ UTR (OR = 0.2, *p* < 0.0001), promoters (OR = 0.3, *p* < 0.0001), and CpG islands (OR = 0.6, *p* < 0.0001) (figure 1 and supplementary table S1). In most of the genomic regions, the majority of DMCs were hypomethylated in the patients compared to controls. In contrast with other genomic regions, in CpG islands, 71% of the DMCs were more methylated in patients than controls.

**Figure 1.**
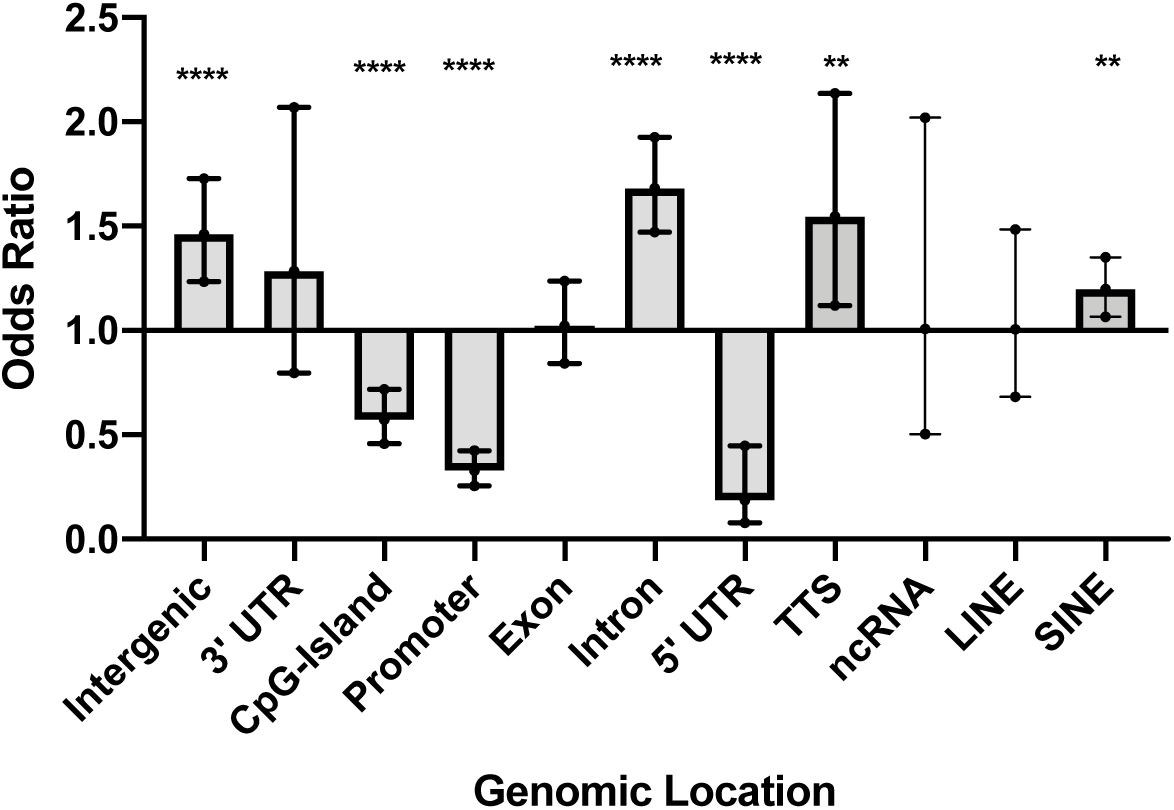
Genomic location of DMC that met an adjusted *p*-value < 0.4. Odds ratio (OR), 95% confidence intervals (CI), and *p*-values were computed against the general distribution of the 3,870,251 CpGs of our dataset using GraphPad Prism. Error bars represent the 95% CI. OR indicate enrichment or depletion of DMCs in each region. transcription termination site (TTS); non-coding RNA (ncRNA); long interspersed nuclear elements (LINE); short interspersed nuclear elements (SINE). \**p*≤0.05, \*\**p*≤0.01,\*\*\**p*≤0.001, \*\*\*\**p*≤0.0001.

The Combined Score approach implemented in RnBeads^29^ was used to identify differentially methylated genes, promoters and CpG islands. A total of 197 (out of 30,771) genes, 112 (out of 29,720) promoters, and 97 (out of 24,117) CpG islands were identified and ranked using this approach. The gene and promoter regions identified are shown in table 2. A total of 9 CpG islands, 17 genes (including RNA, pseudo- and protein-coding genes), and 11 promoters showed significant differential methylation levels between cases and controls at the gene level. The top differentially methylated genes constitute mostly non-coding RNA genes (42%), followed by pseudogenes (27%), then protein-coding genes (19%) (table 2). Among the protein-coding genes, cytidine deaminase (CDA), a marker of monocyte/macrophage differentiation,^34^ is involved in innate immunity pathways. Atypical chemokine receptor 4 (ACKR4) is involved in chemokine signaling.^35^ Distal-less homeobox 5 (DLX5) is a transcription factor involved in bone development and morphogenesis of connective tissue.^36^ The functions of the remaining genes are currently unknown.

**Table 2.** Gene and promoter regions below the Combined Score Cutoff

| Symbol | Gene type | Chr | Position (kb) | MMD | N sites | Rank |
| --- | --- | --- | --- | --- | --- | --- |
| **Genes** |  |  |  |  |  |  |
| RPL30P7 | Pseudogene | 5 | 10489-10489 | 0.31 | 1 | 25 |
| MGC12916 | RNA gene (lncRNA) | 17 | 14207-14209 | 0.23 | 31 | 60 |
| LINC01227 | RNA gene (ncRNA) | 16 | 80601-80607 | -0.21 | 1 | 80 |
| ENSG00000255342 | Uncategorized (lncRNA) | 11 | 123007-123007 | 0.22 | 11 | 81 |
| ENSG00000227930 | RNA gene | 7 | 23931-23937 | -0.2 | 2 | 84 |
| ENSG00000230104 | Uncategorized (lncRNA) | 2 | 173539-173540 | 0.22 | 1 | 131 |
| LOC102724927 | RNA gene (ncRNA) | 16 | 3998-4000 | -0.14 | 5 | 132 |
| ENSG00000229472 | - | 20 | 32669-32670 | 0.18 | 2 | 133 |
| MIR5587 | RNA gene (miRNA) | 16 | 585-585 | 0.22 | 2 | 141 |
| LOC105379365 | RNA gene (ncRNA) | 8 | 34032-34042 | 0.17 | 2 | 146 |
| LOC402634 | Pseudogene | 7 | 2433-2434 | 0.17 | 2 | 163 |
| NEK2P4 | Pseudogene | 2 | 131935-131937 | 0.2 | 7 | 173 |
| NCRNA00250 | RNA gene (ncRNA) | 8 | 135850-135855 | -0.19 | 1 | 180 |
| DLX5 | Protein coding | 7 | 96650-96654 | 0.17 | 114 | 192 |
| LOC101929882 | RNA gene (ncRNA) | 2 | 10179-10181 | 0.13 | 2 | 194 |
| FAM180B | Protein coding | 11 | 47608-47611 | 0.17 | 6 | 195 |
| LOC100652792 | Pseudogene | 15 | 93306-93307 | 0.16 | 4 | 197 |
| **Promoters** |  |  |  |  |  |  |
| CDA | Protein coding | 1 | 20914-20916 | -0.26 | 1 | 7 |
| TAF5LP1 | Pseudogene | 17 | 33824-33826 | -0.22 | 8 | 23 |
| LINC00619 | RNA gene (ncRNA) | 10 | 44339-44341 | -0.25 | 1 | 29 |
| RPL30P7 | Pseudogene | 5 | 10487-10489 | -0.31 | 1 | 45 |
| SNORA25 | RNA gene (snoRNA) | 13 | 106549-106551 | -0.23 | 3 | 72 |
| ENSG00000241456 | RNA gene | 7 | 151123-151125 | 0.17 | 3 | 74 |
| ENSG00000229974 | - | 7 | 134832-134834 | 0.18 | 2 | 94 |
| TMEM140 | Protein coding | 7 | 134831-134833 | 0.18 | 2 | 94 |
| LOC100420018 | Pseudogene | 11 | 35990-35992 | 0.37 | 1 | 101 |
| ACKR4 | Protein coding | 3 | 132315-132317 | -0.23 | 1 | 110 |
| ENSG00000255342 | Uncategorized (lncRNA) | 11 | 123007-123009 | -0.22 | 11 | 112 |

Genes and promoters are ranked based on the RnBeads' Combined Score approach;<sup>29</sup> the rank is computed as the maximum (i.e. worst) of 3 ranks: a) the mean difference in means across all sites in a region of the two groups being compared, b) the mean of quotients in mean methylation, and c) a combined p-value calculated from all site p-values in the region. Chr: chromosome; MMD: Mean Methylation Difference; N: number.

### Gene set enrichment analysis (GSEA)

To gain insight into the most differentially methylated genes and promoters, GSEA^31,37^ was conducted to predict biologically relevant Reactome pathways.^33^ Table 3 lists all Reactome pathways with an FDR ≤ 0.25. This analysis highlighted an immune pathway (immunoregulatory interactions between a lymphoid and a non-lymphoid cell) to be overrepresented in the set of hypermethylated genes, while metabolism pathways (glucuronidation, chondroitin sulfate dermatan sulfate metabolism) showed enrichment among hypomethylated genes. Pathways involved in cell development (regulation of beta cell development and gene expression) and cell signaling (gap junction trafficking, activation of kainate receptors upon glutamate binding, G beta:gamma signalling), were also enriched among hypomethylated genes.

**Table 3.** Summary of Gene Set Enrichment Analysis results

| Entity | Reactome pathway name | Size | ES | NES | p-val | FDR q-val |
| --- | --- | --- | --- | --- | --- | --- |
| Genes | Glucuronidation | 15 | -0.84 | -2.13 | <0.001 | <0.001 |
|  | Chondroitin sulfate dermatan sulfate metabolism | 42 | -0.57 | -1.76 | 0.002 | 0.088 |
|  | Gap junction trafficking | 25 | -0.59 | -1.65 | 0.002 | 0.228 |
|  | G beta: gamma signalling through PI3Kgamma | 24 | -0.59 | -1.65 | 0.003 | 0.191 |
|  | Activation of kainate receptors upon glutamate binding | 29 | -0.55 | -1.60 | 0.008 | 0.236 |
|  | Immunoregulatory interactions between a lymphoid and a non-lymphoid cell | 58 | 0.31 | 1.49 | <0.001 | 0.246 |
| Promoters | Regulation of beta cell development | 27 | -0.64 | -1.92 | <0.001 | 0.022 |
|  | Regulation of gene expression in beta cells | 17 | -0.64 | -1.75 | 0.002 | 0.200 |
|  | Immunoregulatory interactions between a lymphoid and a non-lymphoid cell | 33 | 0.53 | 2.20 | <0.001 | 0.007 |

Pathways with a false discovery rate (FDR) $\leq 0.25$ are shown. Size, number of pathway genes available for analysis; ES, enrichment score for pathway; NES, normalized enrichment score for pathway.

### GO enrichment analysis

To further aid in interpretation of the differentially methylated genes and promoters, we performed enrichment analysis for GO terms associated with the top-ranking genes and promoters in table 4. Multiple development and morphogenesis, immune, and metabolic related terms show enrichment. While hypomethylated genes are enriched for GO terms associated with interferon (IFN) signaling (type I IFN signaling pathway, *p*=8.0E-04; response to type I IFN, *p* = 8.0E-04), hypermethylated genes are enriched for GO terms associated with mesenchyme and epithelial development and cell differentiation (epithelial to mesenchymal transition, *p* = 1.0E-04; nephron tubule formation, *p*=1.0E-04; mesenchymal cell differentiation *p* = 1.0E-04) (table 4).

**Table 4.** Enriched GO terms (*p*≤0.005) among hypo- and hypermethylated regions

| **Table 4.** Enriched GO terms ($p \leq 0.005$) among hypo- and hypermethylated regions | | | |
| --- | --- | --- | --- |
| **ID** | **p-value** | **Term** | **Region** |
| Hypomethylated regions |  |  |  |
| GO:0060337 | 8.00E-04 | type I interferon signaling pathway | genes |
| GO:0034340 | 9.00E-04 | response to type I interferon | genes |
| GO:0070458 | 2.10E-03 | cellular detoxification of nitrogen compound | genes |
| GO:0018916 | 2.80E-03 | nitrobenzene metabolic process | genes |
| GO:0060708 | 2.80E-03 | spongiotrophoblast differentiation | genes |
| GO:0032020 | 4.10E-03 | ISG15-protein conjugation | genes |
| Hypermethylated regions |  |  |  |
| GO:0001837 | 1.00E-04 | epithelial to mesenchymal transition | genes |
| GO:0072079 | 1.00E-04 | nephron tubule formation | genes |
| GO:0048762 | 3.00E-04 | mesenchymal cell differentiation | genes |
| GO:0060980 | 1.00E-03 | cell migration involved in coronary vasculogenesis | genes |
| GO:0048729 | 1.70E-03 | tissue morphogenesis | genes |
| GO:0035295 | 1.70E-03 | tube development | genes |
| GO:0048864 | 1.80E-03 | stem cell development | genes |
| GO:0003218 | 1.90E-03 | cardiac left ventricle formation | genes |
| GO:0070172 | 1.90E-03 | positive regulation of tooth mineralization | genes |
| GO:0072272 | 1.90E-03 | proximal/distal pattern formation involved in metanephric nephron development | genes |
| GO:0072088 | 2.10E-03 | nephron epithelium morphogenesis | genes |
| GO:0061333 | 2.20E-03 | renal tubule morphogenesis | genes |
| GO:0060166 | 2.90E-03 | olfactory pit development | genes |
| GO:0060021 | 2.90E-03 | palate development | genes |
| GO:0060993 | 3.20E-03 | kidney morphogenesis | genes |
| GO:0072080 | 3.20E-03 | nephron tubule development | genes |
| GO:0045893 | 3.50E-03 | positive regulation of transcription, DNA-templated | genes |
| GO:0003166 | 3.90E-03 | bundle of His development | genes |
| GO:0072086 | 3.90E-03 | specification of loop of Henle identity | genes |
| GO:0072513 | 3.90E-03 | positive regulation of secondary heart field cardioblast proliferation | genes |
| GO:2000653 | 3.90E-03 | regulation of genetic imprinting | genes |
| GO:1902680 | 3.90E-03 | positive regulation of RNA biosynthetic process | genes |
| GO:0048598 | 4.20E-03 | embryonic morphogenesis | genes |
| GO:0051891 | 4.80E-03 | positive regulation of cardioblast differentiation | genes |
| GO:0072334 | 1.70E-03 | UDP-galactose transmembrane transport | promoters |
| GO:0035524 | 3.30E-03 | proline transmembrane transport | promoters |
| GO:0060166 | 3.30E-03 | olfactory pit development | promoters |
| GO:2000097 | 3.30E-03 | regulation of smooth muscle cell-matrix adhesion | promoters |
| GO:0001867 | 5.00E-03 | complement activation, lectin pathway | promoters |
| GO:0015820 | 5.00E-03 | leucine transport | promoters |
| GO:0019858 | 5.00E-03 | cytosine metabolic process | promoters |
| GO:0038110 | 5.00E-03 | interleukin-2-mediated signaling pathway | promoters |

Enrichment of Biological Process (BP) Gene Ontology (GO) terms associated with the top-ranking 100 hypo- and hypermethylated genes and promoters as determined by a hypergeometric test implemented in RnBeads.<sup>33</sup>

### Comparison of DNA methylation with previous reports in dermal tissues

The 28 genes and promoters reported in table 2 were compared to results from published genome-wide DNA methylation^14^ and gene expression studies^38-48^ in cultured dermal fibroblasts or skin biopsies. Of our top genes and promoters, two CpGs in *distal-less homeobox 5 (DLX5)* were reported as hypermethylated in skin fibroblasts from dcSSc patients,^14^ which is consistent with our results.

When compared to gene expression profiling studies in cultured dermal fibroblasts or skin biopsies, *DLX5* was reported as under-expressed in patients with SSc,^42^ while *transmembrane protein 140* (*TMEM140*) was reported as overexpressed in patients with SSc,^42^ and correlated with the modified Rodnan skin thickness score (mRSS) in dcSSc patients.^46^

When compared to the genes with compelling evidence of genetic association with SSc,^11^ none of our top 28 genes has been previously reported. Of note, these genome-wide DNA methylation^14^ and gene expression studies^38-48^ in skin-related tissues, as well as genetic association studies,^11^ were all performed in individuals of mostly European ancestry.

### Gene expression of differentially methylated genes

To evaluate the functional effects of DNA methylation on gene expression in our sample of African American subjects, we performed qPCR on the five protein-coding genes (*CDA, TMEM140, ACKR4, DLX5*, *FAM180B*) and three long non-coding (lnc) RNA genes (*MGC12916, LOC102724927, LOC101929882*). These genes were chosen based on their known functions, an increased number of CpG sites detected (>30), and/or detectable transcripts from primary dermal fibroblasts using the RNA isolation/purification technique outlined in the methods section. Of the eight gene transcripts quantified, *DLX5, TMEM140*, and *MCG12916*, showed significant differential expression in cases compared to controls (figure 2). Although these three genes showed hypermethylation, both *DLX5* and *TMEM140* steady state transcript levels were increased, while *MCG12916* steady state transcript levels were decreased in patients compared to controls (figure 2A-C).

**Figure 2.**
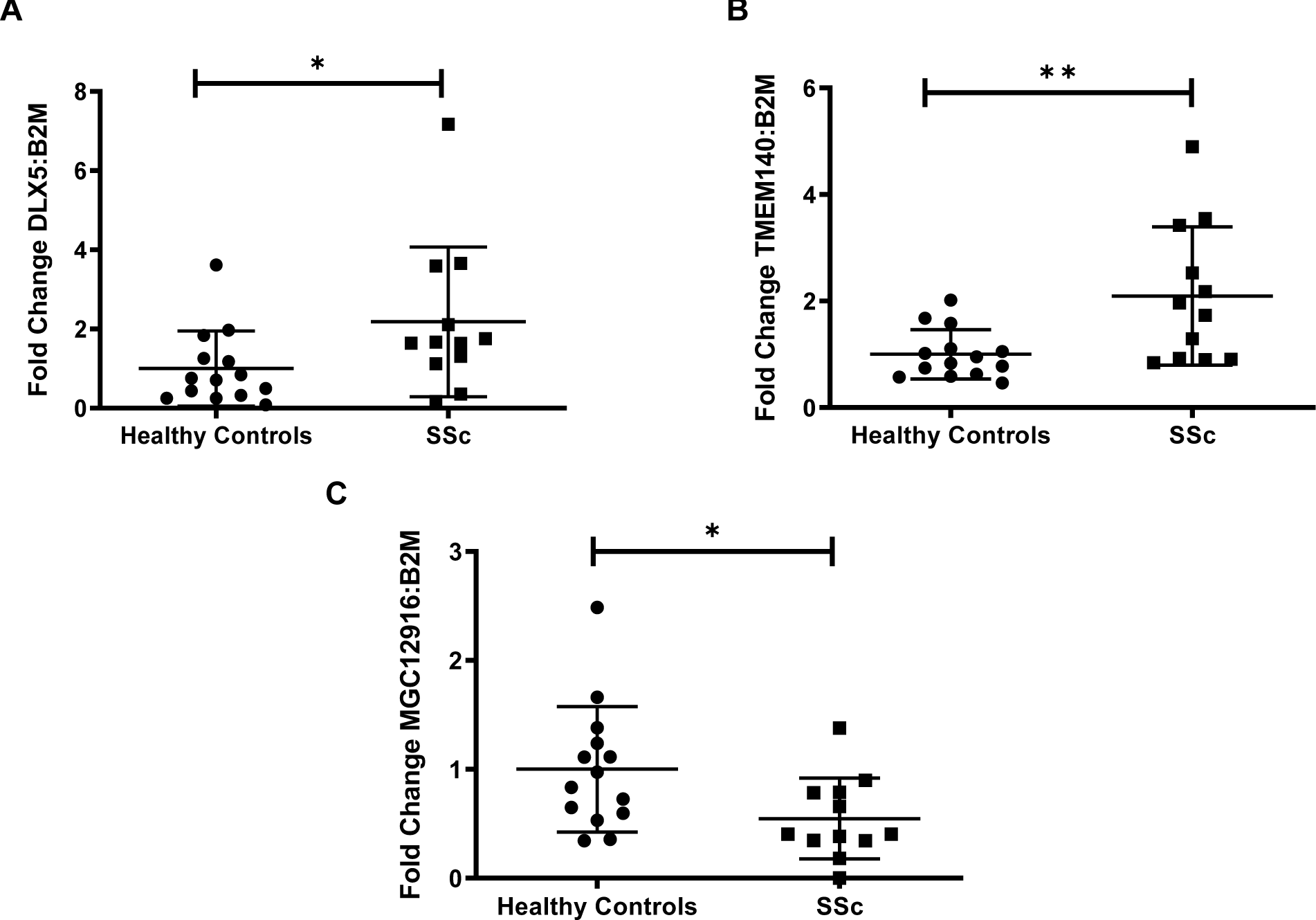
Transcript levels among three differentially expressed genes in AA SSc patients. Among eight genes that were chosen for analysis, three genes demonstrated significantly differentiated expression in AA SSc patients compared to controls. Patient classification detailed on x-axis, while gene transcript level fold change expressed on y-axis. \**p* ≤ 0.05, \*\**p* ≤ 0.01.

## DISCUSSION

This is the first study investigating patterns of differential methylation in primary skin fibroblasts from African American patients with SSc. We found widespread reduced DNA methylation in patients compared with healthy controls, consistent with what has been previously reported in skin fibroblasts from SSc patients of mostly European ancestry,^14^ and peripheral blood from Black South African patients with SSc.^13^

Our findings show novel, top differentially methylated genes constituted mostly of non-coding RNA genes and pseudogenes. Only three protein-coding genes were amongst the top results: *CDA* and *ACKR4* with known roles in immune pathways, and *DLX5* with roles in cell development and proliferation. *DLX5* was previously reported as hypermethylated in skin fibroblasts from dcSSc patients,^14^ which is consistent with our results. However, the previous study analyzed DNA methylation using the HumanMethylation450K array. Our study is based on RRBS which tested eight times more CpGs than those present on the HumanMethylation450K array used in the previous genome-wide study of skin fibroblasts.^14^ Thus, these methods are not directly comparable. Because the array contains only 2% of the CpGs we tested,^49^ minimal overlap can be expected. In addition, extensive differences in DNA methylation are known to exist between individuals of African and European ancestry,^17-24^ due to both variation in genetic ancestry and environmental factors,^19^ with Africans showing higher DNA methylation than Europeans.^20^ These differences help explain the new findings and minimal overlap with previous reports.

*DLX5, TMEM140*, and *MCG12916* exhibited concomitant differential gene expression in the same primary dermal fibroblasts among the differentially methylated genes. While these genes exhibited hypermethylation, *DLX5 and TMEM140* showed overexpression, while *MCG12916* showed downregulation in the same individuals. This is not surprising, as the correlation between DNA methylation and gene expression is positive or negative and is tissue or context specific, in that the local DNA sequence and genomic features largely account for local patterns of methylation.^50-52^ Previous studies report great variation of the quantitative impact of DNA methylation on gene expression among different cell types, with both positive and negative correlations between expression levels and CpG methylation levels.^20,53-56^ Our results show hypomethylation of CpGs was prominent in all regions but CpGs islands, where DMCs were hypermethylated. DMC sites were enriched in introns, intergenic regions, TTS, and SINE, while depleted in 5’ UTR, promoters, and CpG islands. Because CpG sites preferentially located in enhancers are reported to mediate gene expression, not in the promoters, this further supports a modest role of promoters in epigenetic regulatory mechanisms.^20^

Interestingly, despite the differences in tissue and patient characteristics, *TMEM140* was reported as overexpressed in skin biopsy specimens from patients with SSc,^42^ and correlated with the mRSS in dcSSc patients,^57^ which corroborates our findings. On the other hand, *DLX5* was reported as under-expressed in skin biopsy specimens from patients with SSc,^42^ The different outcomes of gene expression for *DLX5* between the experiments could be the result of measuring gene expression in one cell type vs. across multiple cell types in skin biopsies, as well as underlying ancestral differences in gene expression. Although multiple lncRNAs have been reported as dysregulated in SSc patient tissues,^58^ to our knowledge, this is the first report that *MGC12916* has differential gene methylation and expression in primary dermal fibroblasts from African American patients with SSc.

To elucidate the underlying biological processes associated with SSc, GSEA and GO enrichment analyses were conducted. Among the hypomethylated regions, both GSEA and GO enrichment analysis showed enrichment of immune pathways, with GO analysis showing an enrichment in type I IFN signaling. Patients with SSc have excessive IFN and an IFN signature that correlated to early and more severe disease.^59-62^ IFN is also pathogenic in SSc, since exogenous exposure to IFNα or IFNβ leads to its development.^63-66^ The IFN regulatory factor 7 promoter (IRF7) is hypomethylated in SSc peripheral blood mononuclear cells,^67^ supporting the link of IFN signaling and gene hypomethylation in SSc.

Among hypermethylated regions, GSEA showed an enrichment of metabolism, cell development, and cell signaling pathways, and GO enrichment analysis revealed an enrichment in specific pathways related to mesenchyme and epithelial development and cell differentiation. The top enriched GO term among hypermethylated regions, endothelial-mesenchymal cell transition (EMT), is consistent with the current hypothesis that EMT likely influences SSc disease characteristics including endothelial cell dysfunction, dermal fibrosis, and interstitial lung disease.^68,69^ To our knowledge, this is the first reported association between gene and promoter hypermethylation and mesenchymal cell differentiation in SSc. Thus, our GESA and GO analyses correlate with previous data regarding known pathways in SSc.

There are limitations to this study. First, although we are the first to analyze patterns of DNA methylation in dermal fibroblasts in African Americans, the sample size is modest. Nevertheless, with 15 SSc patients, it is comparable to previous genome-wide DNA methylation analyses focused on skin fibroblasts (n = 12 SSc patients),^14^ whole blood (n = 27 SSc patients),^15^ and CD4+ T cells (n = 9 patients),^16^ which included primarily individuals of European ancestry. Second, we were not able to collect a new independent cohort of African Americans to validate the results. Given the rarity of SSc, at the time this study was performed, there were no other comparable prospective studies available. Future studies expanded to multiple centers are needed. Third, and inherent to all epigenomic studies, we cannot exclude the possibility of reverse causation, or whether the DNA methylation changes are a cause or an effect of SSc. Future longitudinal studies will elucidate the role of DNA methylation in disease etiology. Fourth, it is possible that the DNA methylation changes are due to genetic variation. We lack genotypic data on these samples, but note that none of the top differentially methylated genes has been previously reported as associated with SSc. We recognize that it is difficult to account for all lifestyle factors that could affect DNA methylation (i.e. diet, physical activity, body weight, smoking, medications, etc.).^70^ Our samples were balanced relative to smoking and age, so that their confounding effects are minimized. Finally, we do not know the role of *DLX5, TMEM140*, and *MCG12916* in SSc, but we will utilize gene silencing technology to inhibit expression of these genes in primary dermal SSc fibroblasts to elucidate their function in this cell type.

In summary, we identified multiple DNA methylation sites associated with SSc, including sites with evidence of altered methylation in protein-coding, lncRNA and pseudogenes, and concomitant differential expression in *MGC12916, DLX5* and *TMEM140*. Although this cross-sectional study cannot separate causality from response to disease, it identifies DNA methylation alterations in genes and pathways that are important in SSc. Our findings provide a foundation for further research to determine the functional consequences of the differentially methylated loci. Given the reversible nature of epigenetic marks, these loci might represent attractive targets for the treatment or prevention of autoimmune- and/or fibrotic-related diseases.

## Data Availability

Data have been deposited in GEO (Accession code GSE150592). All data are also available from the authors on request.

## Acknowledgements

The authors would like to acknowledge the participants and study coordinator Dr. Carol Lambourne.

## Contributors

PSR designed and coordinated the study. JCO coordinated participant recruitment. JF organized the clinical data. IA and GSB processed the samples and extracted nucleic acids. RCW generated the RRBS data. WS, ESH, PSR and GH analyzed the RRBS data. DBF generated the gene expression data. DBF and CFB analyzed the gene expression data. DBF, KLD and PSR wrote the manuscript. All authors were involved in critical review, editing, revision and approval of the final manuscript.

## Funding

This study was supported by the US National Institute of Arthritis and Musculoskeletal and Skin Diseases of the National Institutes of Health (NIH) under Awards Number T32 AR050958 (DBF), K01 AR067280 (PSR), P30 AR072582 (PSR), R03 AR065801 (PSR), P60 AR062755 (PSR, GSB, JCO), K24 AR060297 (CFB), the MUSC Center for Genomic Medicine, and the National Center for Advancing Translational Sciences of the National Institutes of Health under Award Numbers KL2 TR001452 and UL1 TR001450 (DBF). GH acknowledges support from NIH/NIDA 1U01DA045300-01A1, U54MD010706-CHH and start-up funding from Queens University Belfast.

## Competing financial interests

None declared.

## Patient consent

Obtained.

## Ethics approval

IRB at the Medical University of South Carolina.

## Provenance and peer review

Not commissioned; externally peer reviewed.

## Notes

### Competing Interest Statement

The authors have declared no competing interest.

### Author Declarations

IRB at the Medical University of South Carolina.

